# Tuberculosis in a Brazilian Urban Center: A Decade of Challenges and Epidemiological Trends (2014–2024)

**DOI:** 10.1101/2025.05.01.25326805

**Authors:** Ícaro Raony

## Abstract

Tuberculosis (TB) remains a major public health challenge, particularly in regions with significant social inequalities. This study characterized the epidemiological profile of TB in São José do Rio Preto, a medium-sized city in southeastern Brazil, over an 11-year period. A retrospective analysis of 2,017 cases reported between 2014 and 2024 revealed a predominance among males (male-to-female ratio of 3:1) and adults aged 25–44 years (46.9%), most of whom had not completed high school (82.3%). Pulmonary TB was the most common form (78.5%). High rates of comorbidities were observed, including diabetes (8.5%), alcohol-use disorder (22.8%), smoking (38.7%), illicit drug use (24.1%), and HIV infection (13.8%). HIV co-infection was strongly associated with extrapulmonary TB (OR = 4.3, 99% CI = 3.1–6.2), while TB-related deaths were linked to older age and diabetes (OR = 2.9, 99% CI = 1.5–5.8). Patients undergoing directly observed treatment had markedly less TB-related death (OR = 0.03; 99% CI = 0.0–0.1). These findings highlight that TB in São José do Rio Preto disproportionately affects young adult males with substantial social vulnerabilities, emphasizing the need to address social vulnerabilities and comorbid conditions in TB control efforts.

## 1. Introduction

Tuberculosis (TB) is an infectious and transmissible disease that primarily affects the lungs (pulmonary TB [PTB]), although it can also involve other organs (extrapulmonary TB [EPTB]). The disease is caused by bacteria from the *Mycobacterium tuberculosis* complex and, since 2023, has once again become the leading cause of death from infectious diseases worldwide, surpassing COVID-19 (World Health Organization [WHO], 2024). Coinfection with the human immunodeficiency virus (HIV) remains one of the main factors associated with disease progression, along with alcoholism, smoking, malnutrition, and diabetes (Pai et al., 2016; WHO, 2024). In 2023, it was estimated that approximately 10.8 million new TB cases and 1.25 million TB-related deaths occurred globally (WHO, 2024). In Brazil, specifically, around 103,000 cases and 12,900 deaths were reported during the same year, representing a 15% increase in incidence and an 80% rise in TB-related mortality compared to 2015 (WHO, 2024). Thus, TB continues to be a major public health concern in the country.

Brazil accounts for approximately 50% of the South American territory and is divided into five geographic/administrative regions: North, Northeast, Central-West, Southeast, and South. These regions exhibit distinct geographic, socioeconomic, and political-administrative characteristics, which are reflected in variations in TB incidence, control, and mortality (Cortez et al., 2021). Despite being the most economically developed region, in 2019, the Southeast ranked third in TB incidence and second in TB mortality among Brazil’s regions (dos Santos et al., 2023). In this context, new studies assessing the impact of regional differences on the TB landscape are crucial.

The main objective of the present study was to analyze the epidemiological profile of TB in São José do Rio Preto (SJRP) — a medium-sized municipality located in the state of São Paulo, southeastern Brazil — between 2014 and 2024, as well as to evaluate the impact of socioeconomic factors and comorbidities on TB clinical outcomes, with the aim of providing updated insights to support the development of targeted public health policies.

## 2. Materials and Methods

### 2.1. Ethics Statement

This was a retrospective, descriptive epidemiological study. All data were obtained from the Brazilian Information System for Notifiable Diseases (SINAN, as per its acronym in Portuguese), a publicly accessible government platform. The data are pre-processed by the Brazilian Ministry of Health, including checks for duplicate records, consistency, and completeness (Brazilian Ministry of Health, 2020). In accordance with Resolution No. 466/12 of the Brazilian National Health Council, this study was exempt from approval by a research ethics committee, as it used only secondary, public, and anonymized data.

### 2.2. Study Area and Target Population

The municipality of SJRP is located in the northern part of São Paulo state, approximately 480 km from the state capital. Classified as a medium-sized municipality, SJRP has a population of 480,393 inhabitants according to the latest census by the Brazilian Institute of Geography and Statistics (IBGE, 2022), with a population density of 1,111.17 inhabitants/km^2^. Approximately 95.8% of households have access to adequate sanitation services, and the municipality presents a high Municipal Human Development Index (MHDI) of 0.797 (based on 2010 data), ranking 50th nationally (IBGE, 2022). Nevertheless, about one-third of the population lives with a monthly per capita income below half the minimum wage (IBGE, 2022), highlighting persistent social inequalities in the city.

### 2.3. Data Collection and Variables

All reported cases of TB in SJRP from January 1, 2014, to December 31, 2024, were extracted from the SINAN database (accessed on April 26, 2025). For analysis, the annual number of TB cases and TB-related deaths were stratified by the following sociodemographic and clinical variables: sex (male, female); age group (0–14, 15–24, 25–34, 35–44, 45–54, 55–64, ≥65 years); skin color (yellow, white, indigenous, brown, black, missing information); education level (completed higher education, completed secondary education, completed primary education, incomplete primary education, illiterate, not applicable, missing information); clinical form of TB (PTB, EPTB, PTB + EPTB); specific extrapulmonary manifestations (pleural, peripheral lymphatic, osseous, ocular, miliary, meningoencephalic, others); result of the rapid molecular test for TB (RMT-TB: positive rifampicin-sensitive, positive rifampicin-resistant, negative, inconclusive, not performed, missing information); performance of directly observed therapy (DOT: yes, no, missing information); and case outcome (cure, death from TB, death from other causes, treatment abandonment, primary abandonment, transfer, drug-resistant TB [DR-TB], treatment regimen change, missing information). In addition, risk factors associated with TB cases and deaths were evaluated, including smoking, alcohol-use disorder, illicit drug use, and diabetes.

### 2.4. Statistical Analysis

Statistical analyses were performed using GraphPad Prism software, version 10.0. Variables were presented as absolute numbers or percentages, as indicated in tables and figures. Categorical trend analyses were conducted using the two-tailed Fisher’s exact test, combined with the Baptista-Pike method to estimate odds ratios (ORs) and their corresponding confidence intervals (CIs). A significance level of *p* < 0.01 was adopted for all statistical analyses.

## 3. Results

Between January 1, 2014, and December 31, 2024, a total of 2,017 cases of TB were reported in SJRP, with an average of 183.4 cases per year (99% CI = 152.9–213.8) (**Figure 1a**). Most patients were male (75.2%), white (60.8%), and had not completed secondary education (82.3%). Adults aged 25–44 years accounted for 46.9% of all cases (**Figures 1a–c**; **Table I**). Among vulnerable populations, 299 cases (14.8%) occurred in people deprived of liberty (PDL), 59 cases (2.9%) in individuals experiencing homelessness, and 42 cases (2.1%) in healthcare workers (**Table I**).

**Table I.**
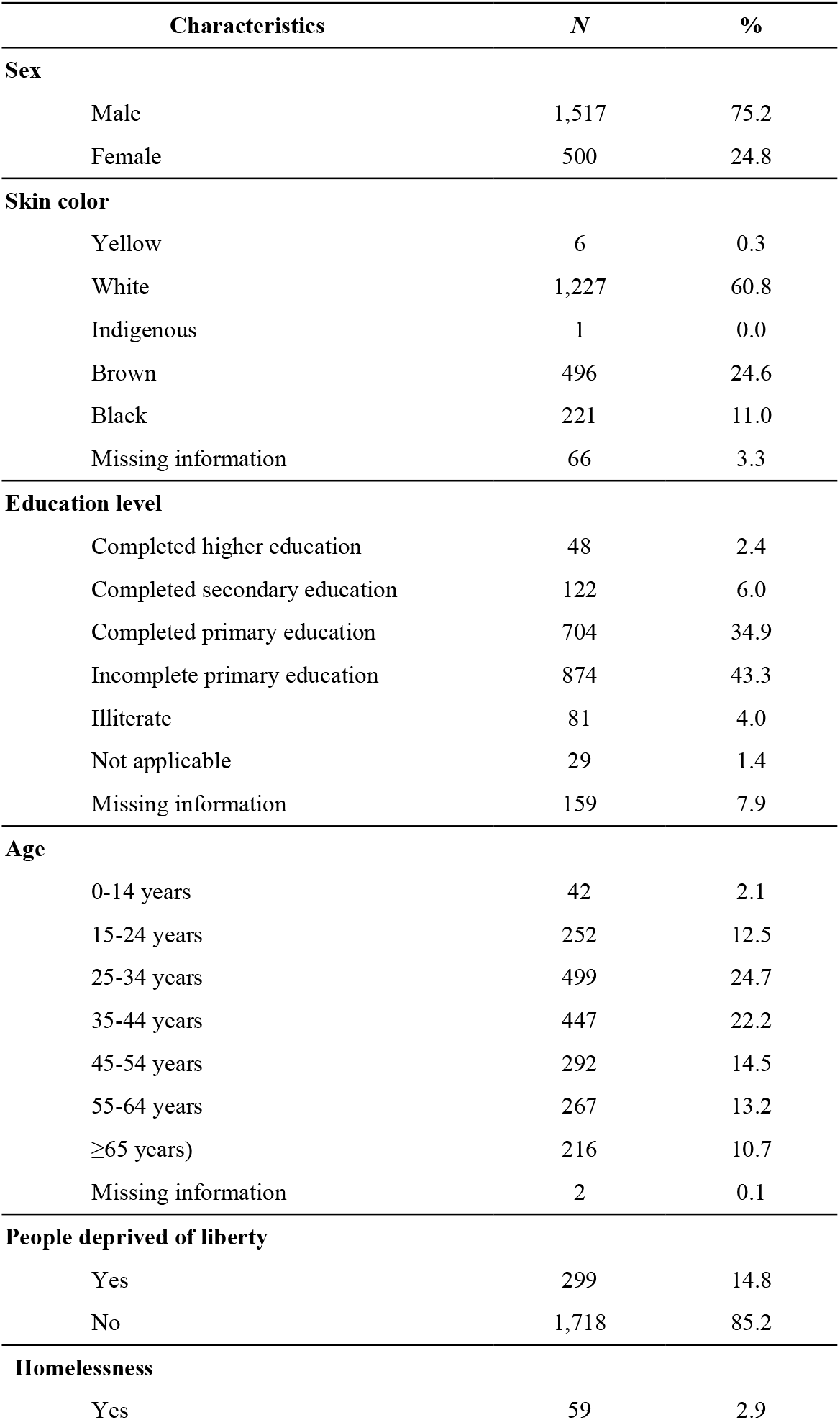
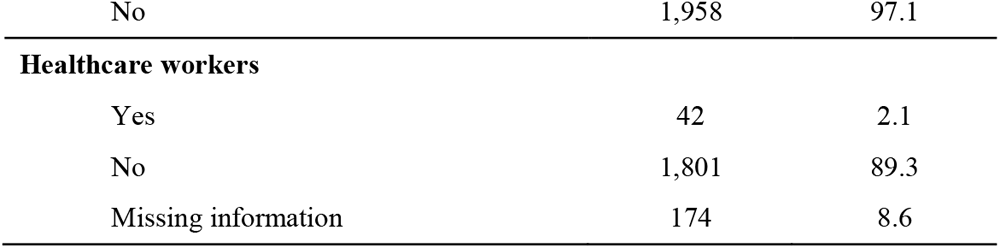
Sociodemographic characteristics of patients diagnosed with TB in SJRP from 2014 to 2024 (*N* = 2,017).

**Figure 1.**
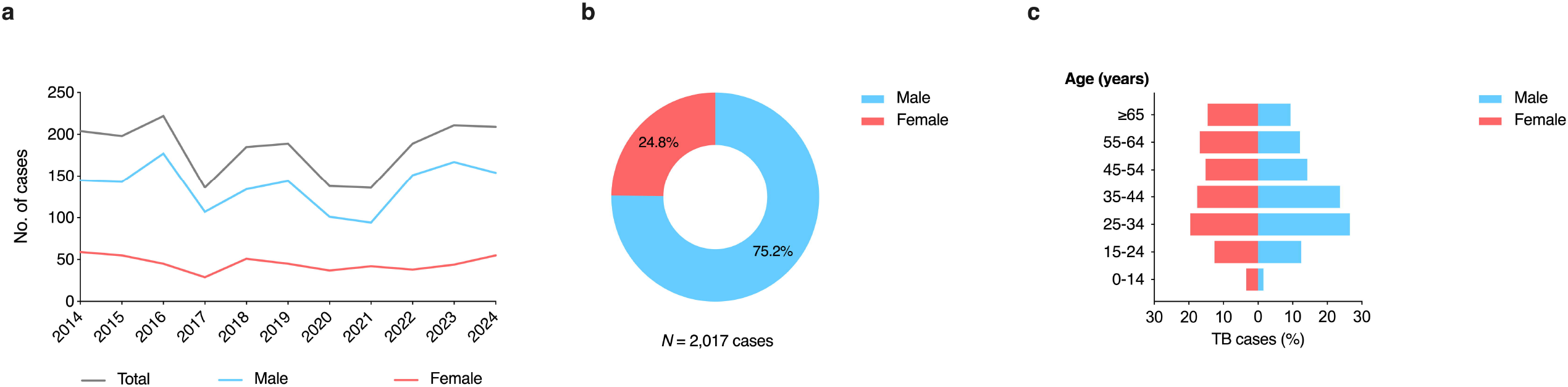
Distribution of TB cases by sex and age group in SJRP, between 2014 and 2024. (**a**) Annual distribution of TB cases by sex. (**b**) Overall proportion of TB cases by sex during the study period. (**c**) Distribution of TB cases by sex and age group, expressed as a percentage of the total cases for each sex.

The most prevalent clinical form was isolated PTB, accounting for 78.5% of cases. Exclusively EPTB represented 14.6% of cases, while mixed forms (PTB + EPTB) comprised 6.9% (**Figure 2a**; **Table II**). Among extrapulmonary manifestations, miliary TB was the most common (42.9%), followed by pleural (7.1%) and osseous forms (7.1%) (**Figure 2b**). Regarding associated comorbidities, 8.5% of patients had diabetes, 22.8% reported alcohol-use disorder, 38.7% were smokers, 24.1% reported illicit drug use, and 13.8% were people living with HIV (PLHIV) (**Table II**). A positive association was observed between HIV coinfection and extrapulmonary TB manifestations (OR = 4.3; 99%, CI = 3.1–6.2, *p* < 0.0001).

**Table II.** Clinical characteristics of patients diagnosed with TB in SJRP from 2014 to 2024 *(N =* 2,017).

| Characteristics | N | % |
| --- | --- | --- |
| **Clinical form** |  |  |
| Pulmonary | 1,583 | 78.5 |
| Extrapulmonary | 294 | 14.6 |
| Pulmonary + Extrapulmonary | 140 | 6.9 |
| **Diabetes** |  |  |
| Yes | 172 | 8.5 |
| No | 1,845 | 91.5 |
| **Alcohol-use disorder** |  |  |
| Yes | 460 | 22.8 |
| No | 1,557 | 77.2 |
| **Smoking** |  |  |
| Yes | 781 | 38.7 |
| No | 1,236 | 61.3 |
| **Illicity drug use** |  |  |
| Yes | 487 | 24.1 |
| No | 1,530 | 75.9 |
| **People living with HIV** |  |  |
| Yes | 279 | 13.8 |
| No | 1,738 | 86.2 |
| **Rapid molecular test for TB** |  |  |
| Positive rifampicin-sensitive | 547 | 27.1 |
| Positive rifampicin-resistant | 36 | 1.8 |
| Negative | 179 | 8.9 |
| Inconclusive | 26 | 1.3 |
| Not performed | 1087 | 53.9 |
| Missing information | 142 | 7.0 |
| **Directly observed therapy** |  |  |
| Yes | 864 | 42.8 |
| No | 1,036 | 51.4 |
| Missing information | 117 | 5.8 |
| **Case outcomes** |  |  |
| Cure | 1,371 | 68.0 |
| TB-related death | 94 | 4.7 |
| Death from other causes | 149 | 7.4 |
| Treatment abandonment | 207 | 10.3 |
| Primary abandonment | 3 | 0.1 |
| Transfer | 26 | 1.3 |
| Drug-resistant TB | 23 | 1.1 |
| Treatment regimen change | 16 | 0.8 |
| Missing information | 128 | 6.3 |

**Figure 2.**
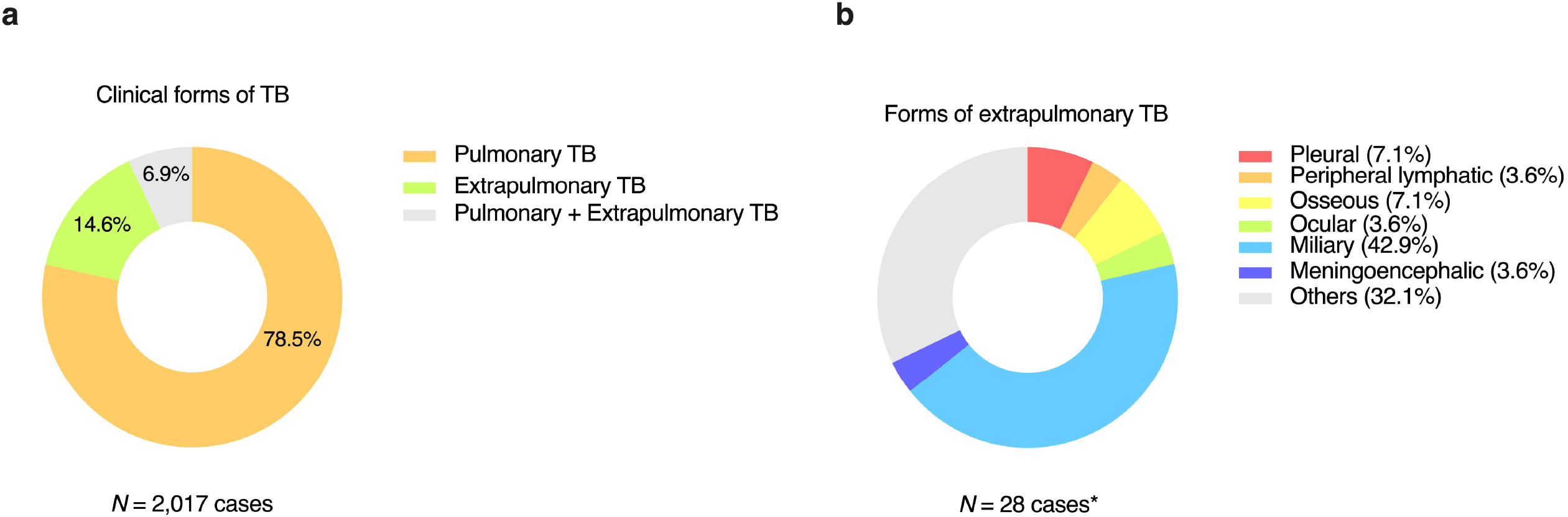
Clinical forms of TB in SJRP, between 2014 and 2024. (a) Frequency of TB clinical forms: exclusively PTB (*N* = 1,583; 78.5%), exclusively EPTB (*N* = 294; 14.6%), and mixed (PTB + EPTB) (*N* = 140; 6.9%). (b) Frequency of EPTB manifestations. ^*^Of the 434 cases involving extrapulmonary sites, 406 were excluded from this analysis due to missing information on the affected site in the SINAN database.

Among the 2,017 reported cases, the RMT-TB was not performed in 53.9% of patients, and 7.0% had missing data (**Table II**). Thus, among the 788 cases with available RMT-TB results, 69.4% were rifampicin-sensitive and 4.6% showed rifampicin resistance. Regarding DOT, among patients with available information (n = 1,900), 45.5% underwent supervised treatment (**Table II**). Concerning clinical outcomes, among cases with available information (n = 1,889), 72.6% achieved cure, while 11% discontinued treatment, including three cases classified as primary abandonment. TB-related deaths accounted for 5% of cases, and 7.9% of cases resulted in death from other causes (**Figure 3a**; **Table II**).

**Figure 3.**
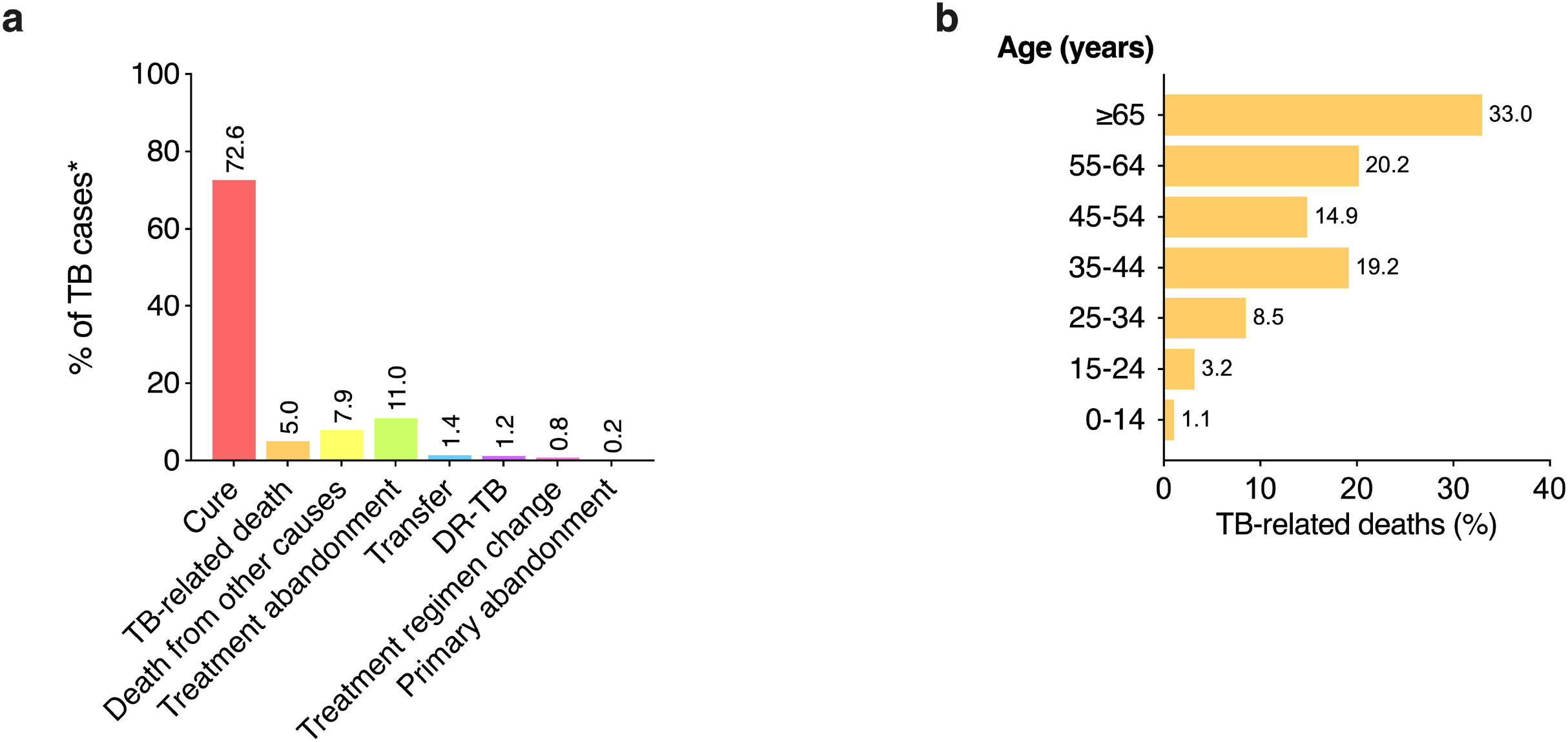
Clinical outcomes of TB cases and distribution of TB-related deaths by age group in SJRP, between 2014 and 2024. (**a**) Distribution of TB cases according to clinical outcomes: cure, TB-related death, death from other causes, treatment abandonment, primary abandonment, transfer, development of DR-TB, or treatment regimen change. ^*^Of the 2,017 cases, 128 were excluded from this analysis due to missing outcome information (*N* = 1,889 cases). (b) Percentage distribution of TB-related deaths (*N* = 94) by age group.

There was a progressive increase in the frequency of TB-related deaths with advancing age, with one-third of deaths occurring in individuals aged 65 years or older (**Figure 3b**). Patients with diabetes had nearly three times the odds of TB-related death compared to non-diabetic patients (OR = 2.9; 99% CI = 1.5–5.8; *p* = 0.0002). Conversely, DOT showed a significant protective effect, reducing the likelihood of TB-related death by 33-fold compared to patients who did not undergo DOT (OR = 0.03; 99% CI = 0.0–0.1; *p* < 0.0001) (**Table III**). No significant association was observed between TB-related death and sex, skin color, alcohol-use disorder, smoking, or illicit drug use (**Table III**).

**Table III.** Association between sociodemographic and clinical variables with TB-related deaths in SJRP from 2014 to 2024.

| Variables | OR | 99% IC | *p* |
| --- | --- | --- | --- |
| **Sex** |  |  |  |
| Male vs. female | 1.1 | 0.6-2.1 | 0.8077 |
| **Skin color** |  |  |  |
| White vs. non-white | 1.9 | 1.0-3.6 | 0.0102 |
| **Directly observed therapy** |  |  |  |
| Yes vs. no | 0.03 | 0.0-0.1 | <0.0001 |
| **Comorbidities (yes vs. no)** |  |  |  |
| Diabetes | 2.9 | 1.5-5.8 | 0.002 |
| Alcohol-use disorder | 1.4 | 0.9-2.2 | 0.1667 |
| Smoking | 1.0 | 0.6-1.8 | 0.9139 |
| Illicit drug use | 0.5 | 0.2-1.1 | 0.0352 |

Odds ratios (OR) and corresponding 99% confidence intervals (CI) for TB-related death according to sociodemographic and clinical characteristics, estimated using the two-tailed Fisher's exact test combined with the Baptista-Pike method.

## 4. Discussion

TB remains one of the leading causes of morbidity and mortality worldwide, with a particularly high burden in regions marked by social inequality, as observed in SJRP, a medium-sized municipality in São Paulo state. Between 2014 and 2024, the municipality reported 2,017 TB cases, with an average of 183.4 cases per year. The predominance of cases among young adult males and the high prevalence of comorbidities, such as smoking, alcoholism, and diabetes, reflects an epidemiological profile similar to that observed in other regions of Brazil and worldwide (WHO, 2024).

Nationally, the incidence of TB in the Southeast region of Brazil is relatively high, with the Brazilian Southeast region - encompassing the states of Espírito Santo, Minas Gerais, Rio de Janeiro and São Paulo - presenting the highest TB notification rate (Meirelles and Palha, 2019). Although São Paulo is considered one of the most developed states, socioeconomic and behavioral factors, such as inequality and the high prevalence of comorbidities, continue to impact the disease burden. In the present study, approximately 14.8% of TB cases occurred among incarcerated individuals, aligning with national literature that highlights the high prevalence of TB in this vulnerable population (Cords et al., 2021). HIV coinfection was also an important factor, with 13.8% of TB occurring in PLHIV. A significant association between HIV coinfection and extrapulmonary TB manifestations was also observed, supporting previous studies indicating HIV coinfection as one of the main risk factors for disseminated and EPTB, which is often associated with CD4 lymphopenia (Meintjes and Maartens, 2024; Tornheim and Dooley, 2017).

The predominance of PTB among SJRP patients is consistent with global trends, where pulmonary remains the most common form of TB, followed by EPTB, which represents a smaller proportion of cases (Zumla et al., 2013). The present findings indicates that the miliary form was the most frequent extrapulmonary manifestation in SJRP, suggesting a clinically relevant pattern that should be monitored, as miliary TB is typically associated with a poorer prognosis, as well as more challenging diagnosis and treatment (Sharma et al., 2016).

The association between diabetes and TB-related death observed in the present study is consistent with international data showing that patients with diabetes are at increased risk of developing severe forms of TB and experiencing poorer clinical outcomes (reviewed by Boadu et al., 2024). Diabetes not only predisposes individuals to greater susceptibility to TB infection but also exacerbates the severity of the disease, potentially due to compromised immune responses, particularly when diabetes is poorly controlled (Boadu et al., 2024). This underscores the importance of policies targeted at reducing the burden of diabetes are needed to contribute to the aims of ending TB.

The percentage of SJRP patients who died due to TB was 5%, which is concerning, given the potential for reducing this rate through effective interventions such as DOT, which involves observing a patient swallow their TB medications to increase the individual’s adherence therapy. DOT was highly protective in the present study, reducing the likelihood of TB-related death by a factor of 33. Despite the national recommendation for all individuals diagnosed with TB to undergo DOT, the present results indicates that less than half of the patients in SJRP from 2014 to 2024 received DOT, which corroborates findings from other Brazilian studies (Reis-Santos et al., 2015; Soares et al., 2006). Therefore, the current data reinforces the importance of DOT as a cost-effective strategy to improve clinical outcomes and reduce TB-related death in Brazil.

Globally, TB remains a significant public health challenge, especially in developing countries. In conclusion, this study presents a concerning yet insightful picture, providing critical insights for the planning and implementation of effective TB control policies, particularly for socially vulnerable populations. Public health strategies must prioritize addressing the specific needs of at-risk groups, such as individuals with diabetes and substance users. By adopting integrated approaches that consider both the social determinants and clinical context of patients, we can more effectively address the ongoing challenges of TB in SJRP, Brazil and worldwide.

## Data Availability

All data produced in the present work are contained in the manuscript.

## Acknowledgments

This study was indirectly supported by fellowship granted by Coordenação de Aperfeiçoamento de Pessoal de Nível Superior, (CAPES/Brazil; finance code 001).

## List of Abbreviations

CI: (confidence interval)
DOT: (directly observed therapy)
DR-TB: (drug-resistant tuberculosis)
EPTB: (extrapulmonary tuberculosis)
HIV: (human immunodeficiency virus)
IBGE: (Brazilian Institute of Geography and Statistics)
MHDI: (Municipal Human Development Index)
OR: (odds ratio)
PDL: (people deprived of liberty)
PLHIV: (people living with HIV)
PTB: (pulmonary tuberculosis)
SINAN: (Brazilian Information System for Notifiable Diseases)
RMT-TB: (rapid molecular test for tuberculosis)
SJRP: (São José do Rio Preto)
TB: (tuberculosis)
WHO: (World Health Organization)

